# Systematic computational fluid dynamic analysis of intra-aneurysmal blood flow using data-driven synthetic cerebral aneurysm geometries

**DOI:** 10.64898/2026.02.28.26347304

**Authors:** Yasuhiro Yamamoto, Kakeru Ueda, Hiro Wakimura, Shigeki Yamada, Yoshiyuki Watanabe, Hiroto Kawano, Satoshi Ii

## Abstract

The present study presents a systematic approach for generating data-driven synthetic cerebral aneurysm geometries and evaluating their hemodynamics through computational fluid dynamics. Seven patient-specific aneurysm geometries from the right internal carotid artery were reconstructed from time-of-flight magnetic resonance angiography images and standardized through orientation alignment, followed by non-rigid registration onto a common spherical point cloud as a template. Principal component analysis (PCA) was then applied to the aligned point-cloud data to quantify morphological variability and parameterize shape deformation. The first four principal components captured over 90% of the total variance; however, higher-order components were required to capture the detailed geometrical features of the original geometries. Computational fluid dynamic simulations were performed on the PCA-based synthetic geometries under pulsatile flow conditions to investigate the influence of shape variations on intra-aneurysmal flow patterns, time-averaged wall shear stress (TAWSS), and oscillatory shear index (OSI). The first principal component score (PCS1), which was associated with changes in aneurysm height and dome width, had the strongest effects on TAWSS and OSI levels. Lower PCS1 values, which corresponded to taller and more oblique domes, produced slower adjacent flow and elevated OSI, whereas higher PCS1 values increased TAWSS. The second principal component score primarily modulated lateral geometric asymmetry and further influenced OSI distribution for the lower PCS1 values. Collectively, these findings indicate that PCA-based shape parameterization provides a practical approach for generating synthetic aneurysm datasets and systematically assessing how specific morphological features govern hemodynamic behavior. The proposed approach is expected to contribute to the future development of surrogate modeling and data-driven hemodynamic prediction.

## 1. Introduction

Subarachnoid hemorrhage carries a high mortality rate (approximately 35%); its major cause is the rupture of cerebral aneurysms (Donkor, 2018). Aneurysms are detected in approximately 2% of adults without risk factors for subarachnoid hemorrhage, and the annual risk of rupture in these cases remains relatively low, at approximately 0.7% (Rinkel et al., 1998). Surgical treatments, such as clipping via craniotomy and coil embolization using catheters, are commonly performed; however, these procedures entail risks of complications and mortality (Sharma et al., 2019). It is therefore clinically important to screen high-risk patients who have asymptomatic aneurysms that were discovered incidentally. The growth and rupture of cerebral aneurysms are influenced by aneurysm morphology and intra-aneurysmal hemodynamics (Meng et al., 2014; Penn et al., 2011; Signorelli et al., 2018). Various indices related to wall shear stress (WSS) have been investigated in previous studies (Staarmann et al., 2019; Sun et al., 2020).

Many studies have explored the diverse morphological characteristics of aneurysms to clarify their relationship with rupture (Sanchez et al., 2023). Juchler et al. (2022) evaluated various shape parameters, including asphericity and curvature, using a publicly available dataset of ruptured and unruptured aneurysms. Eulzer and Lawonn (2025) proposed a parametric mapping method to represent aneurysm shapes using multiple public datasets and investigated how each principal mode in a principal component analysis (PCA) contributed to rupture status. The findings of these studies suggest that not only aneurysm size but also morphological features are associated with rupture. Elucidating the relationship between these morphological features and aneurysmal hemodynamics is of clinical importance.

Computational fluid dynamic (CFD) analysis using patient-specific data has become the standard tool for evaluating intra-aneurysmal blood flow and related hemodynamic factors (Ii et al., 2018; Wu et al., 2022; Ichimura et al., 2024), and efforts have been made to connect these findings to rupture risk (Valen-Sendstad et al., 2018; Murayama et al., 2019; Bozorgpour, 2026). Readers may refer to recent comprehensive reviews on this topic (Liang et al., 2019; Loly et al., 2025). In such analyses, vascular geometry is reconstructed from patient-specific image data, and numerical simulations are performed. However, given the limited amount of clinical data that are available, it is challenging to establish universal relationships between diverse morphological features and hemodynamic factors. Consequently, CFD analyses using synthetic geometries have recently gained attention. Although they did not focus on cerebral aneurysms, several studies have applied PCA to multiple real vascular geometries to construct statistical shape models, generated synthetic vessels by varying each mode, and conducted CFD to explore the relationship between morphological features and hemodynamics (Du et al., 2022; Wiputra et al., 2023). Other studies have used manually deformed synthetic shapes (Faisal et al., 2025) or synthetic geometries to simulate aneurysm development (Li et al., 2025). Nevertheless, a systematic method for generating synthetic geometries and conducting CFD specifically for cerebral aneurysms has not yet been established.

The aim of the present study was to propose a systematic CFD approach based on data-driven synthetic geometries for evaluating hemodynamics in cerebral aneurysms. In this approach, point-cloud data of aneurysm shapes obtained from patient-specific data were registered to a template using non-rigid registration. PCA was then applied to the template point cloud. By changing the scores of principal components based on the resulting principal vectors, multiple aneurysm geometries that were consistent with real geometries were generated. Using these synthetic geometries, CFD analyses were performed to clarify how variations in major principal components correspond to changes in intra-aneurysmal flow characteristics and WSS.

## 2. Material and methods

### 2.1 Reconstruction of patient-specific aneurysm geometry

We used time-of-flight magnetic resonance angiography images from seven patients with cerebral aneurysms in the right internal carotid artery. The data acquisition was approved by the Ethics Committee of Shiga Medical University (IRB Number: R2019-227). The study was conducted according to the approved guidelines of the Declaration of Helsinki. Each time-of-flight magnetic resonance angiography image had a pixel size of 0.3906 mm and a slice thickness of 0.6 mm. Geometries were extracted using the medical imaging software Mimics (Materialise, Leuven, Belgium). Figure 1 shows the reconstructed vessel geometries.

**Fig. 1.**
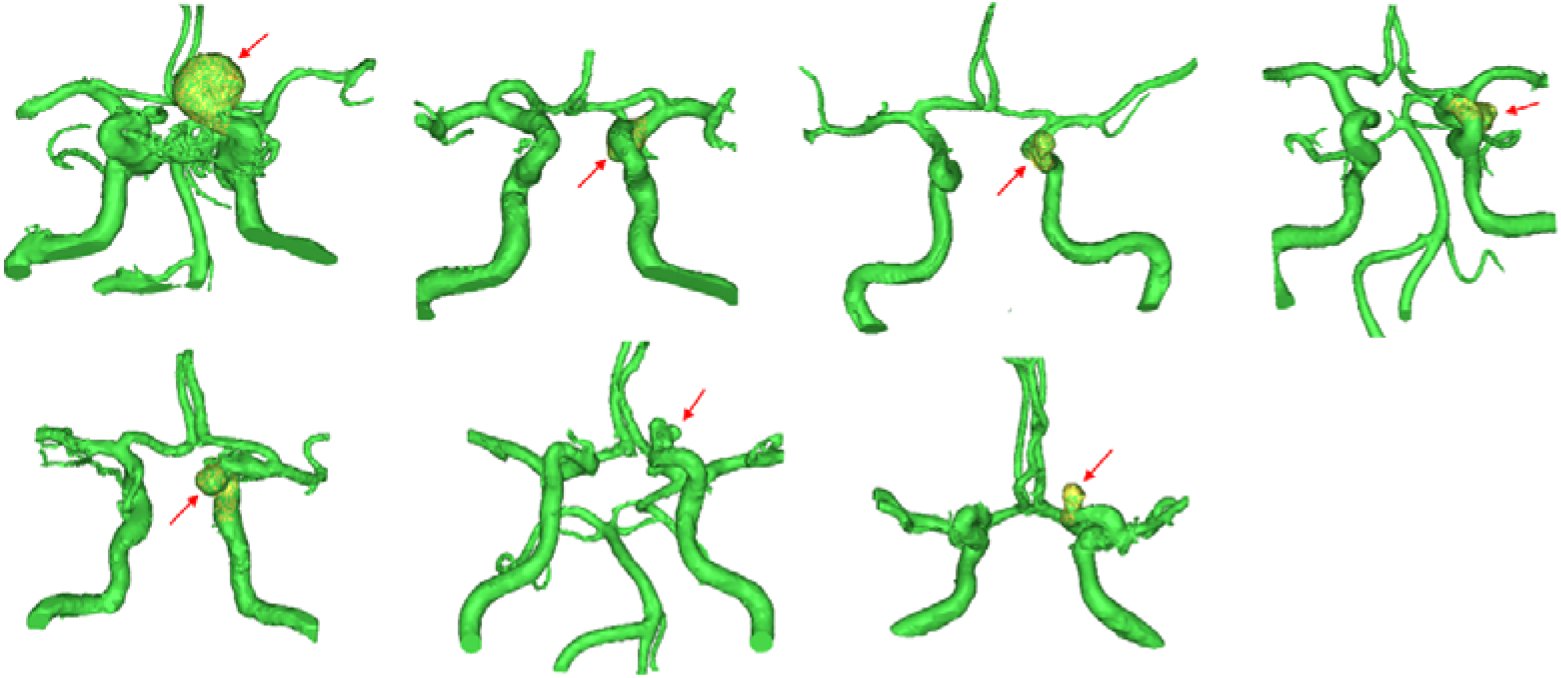
Reconstructed vessel geometries from patient-specific time-of-flight magnetic resonance angiography images, where the red arrow denotes the aneurysm region.

In the present study, to reduce the complexity of shape diversity, we focused our analysis on aneurysms only. Aneurysm geometry was manually extracted from the reconstructed vessel using Meshmixer (Autodesk, San Francisco, CA, USA), and the cutting plane around the aneurysm neck was manually determined. We labeled each aneurysm as sample 1–7 (S1–S7) (Fig. 2(A)).

**Fig. 2.**
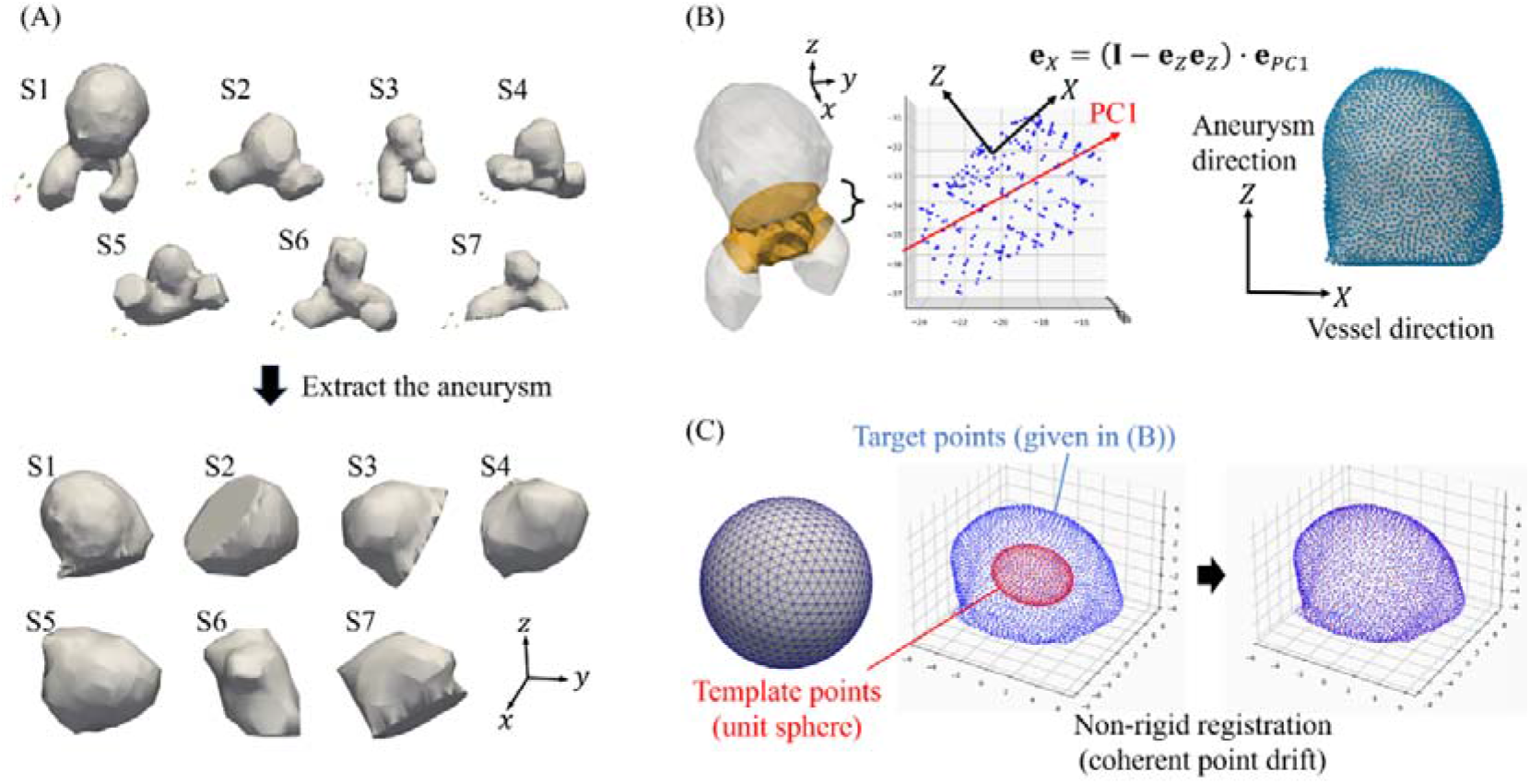
Preprocessing of morphological geometries of cerebral aneurysms prior to principal component analysis (PCA). (A) Extraction of the cerebral aneurysms. (B) Orientation alignment of the cerebral aneurysms. (C) Coherent point drift–based non-rigid registration of aneurysm shape using a spherical pointset as the template.

The orientation of an aneurysm against the main vessel is an important process in evaluating hemodynamics because the flow circulation inside the aneurysm is highly affected by inlet flow around the aneurysm neck. Thus, we adjusted the orientations of the aneurysms for all samples by taking a rigid rotation from the global coordinate system (*x, y, z*) to an orthogonal coordinate system (*X, Y, Z*), where the components showed the axes of the vessel direction, span direction in blood flow, and aneurysm direction. The unit basis vector for the aneurysm axis, **e**_*Z*_, was set to the unit normal vector of the aneurysmal cutting–plane for the neck, whereas the vessel axis *X* was systematically calculated using PCA for a main vessel geometry excluding the aneurysm region. Equation (1) shows the data matrix **A** ∈ ℝ ^*n*× 3^ that was used in the PCA, which consisted of coordinate points of the vessel surface (*x*_*i*_, *y*_*i*_, *z*_*i*_) for *i* ∈ [**1**,*n*]. Next, the data matrix 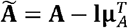 was introduced, where was introduced, where I =(1,1,1,…]^*T*^ ∈ ℝ^*n*^ and ***μ***_*A*_ =(*μ*_*A1*_, *μ*_*A2*_, *μ*_*A3*_,…)^*T*^ ∈ ℝ^*n*^ were the column vectors consisting of unity and the mean value *μ*_*Ai*_ of each column *i* of **A**. The evaluated first principal component, **e**_*PC*1_, was then expected to describe the vessel direction. Because there was no guarantee that was **e**_*PC*1_ perpendicular to **e**_*Z*_, we finally calculated the unit basis vector of the vessel direction, **e**_*X*_, by taking a surface projection to the perpendicular plane of the aneurysm axis as **e**_*X*_ =(**I** − **e**_*Z*_ **e**_*Z*_) · **e**_*PC*1_, where **I** was the identity tensor (Fig. 2 (B)).

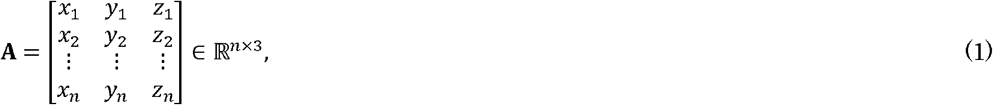

Although the PCA requires a common metric for representing geometry among samples, the surface points of the extracted aneurysms are independent among samples. To address this issue, we introduced a template and fitted the coordinate points of the template to those of the reconstructed aneurysm using a non-rigid registration based on coherent point drift (CPD) (Myronenko and Song 2021; Gatti et al., 2022). The template (or reference) was given as nodes on a spherical mesh that was based on a geodesic regular icosahedron with 1002 vertices, and the target was set to the coordinate points of the smoothed geometry of the reconstructed aneurysm (Fig. 2(C)). The parameters in CPD were as follows: *β* = 2 for the spatial scale of deformation; *λ* =3 for the regularization strength of deformation; *tol* = 10^−5^ for the convergence threshold; and *N*_*ite*_ =50 for the maximum iterations.

### 2.2 PCA for patient-specific aneurysm geometries

In the present study, nondimensional governing equations were used in fluid simulations, meaning that each aneurysm size emerged in the Reynolds number as a representative length. For this reason, each aneurysm size was normalized so that a radius of an equivalent sphere with the same volume of the aneurysm became the unity.

Point-cloud data of the CPD-based aneurysm geometries in Section 2.1 were used for PCA. The data matrix **B** was defined as equation (2), where 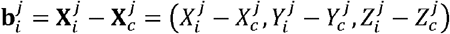 was the *i*-th coordinate point vector in the local coordinate system (*i* ∈ [1,*N*_*v*_ = 1002]) from each centroid 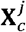 for the sample *j* ∈ [1,*N*_*s*_ = 7]. The PCA was applied for the modified matrix 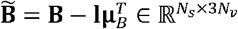, which was the deviation that was calculated by subtracting the mean value of each column 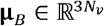. This indicates that the PCA was performed for directional vectors at *N*_*v*_ points from the average geometry for *N*_*s*_ samples.

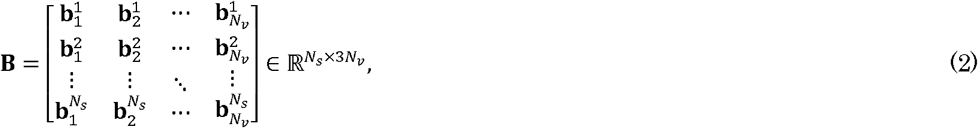

### 2.3 Generation of data-driven synthetic geometries (PCA-based aneurysm geometries)

Arbitrary geometries were generated by calculating a principal component score (PCS) for the PCA of patient-specific geometries in Section 2.2. Through the PCA, a singular value decomposition was performed as 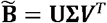, where 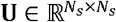 and 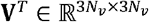 ^V^ were the square matrices and 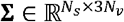, was the matrix whose diagonal components were given by singular values *σ*_*k*_ (*k* ∈ [1,*N*_*σ*_]) with *N*_*σ*_ = min (*N*_*s*_,3 *N*_*v*_), and non-diagonal components were given by 0. Because of the features of singular value decomposition, a set of directional vectors for *j*-th sample 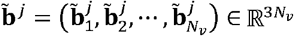, was represented by

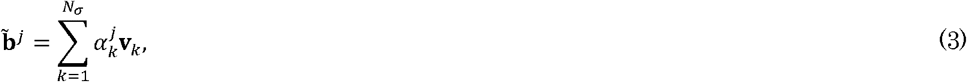

where 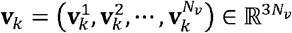 ^V^ was the *k*-th unit basis vector in the singular value decomposition (*k* ∈ [1, *N*_*σ*_]) as the *k*-th column vector of **V**, and 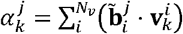 was the *k*-th PCS for *j*-th sample. The arbitrary directional vector was then generated by

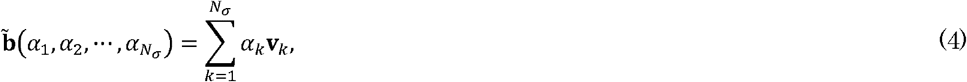

where *α*_*k*_ was the component of the unit basis *v*_*k*_ (*k* ∈ [1, *N*_*σ*_]), which can be considered as an arbitrary PCS to generate an aneurysm geometry based on the PCA. Subsequently, an arbitrary aneurysm geometry was generated by adding the obtained directional vectors 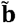 to the average geometry 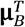. Importantly, in the view point of model order reduction, the geometry can be approximated by *N*_*σ*_ being replaced by *M*_*PC*_ (≤ *N*_*σ*_) in equations (3) and (4).

For CFD analyses, we aligned all centroids of the synthetic geometries to the origin and re-scaled the size so that the radius of an equivalent sphere (with the same volume) was unity. Given that the bottom surface of each aneurysm was no longer planar, we excluded the bottom region of the generated aneurysms below a plane that was orthogonal to the aneurysm direction **e**_*Z*_, on the same *Z*-position.

### 2.4 CFD using PCA-based aneurysm geometries

CFD analyses were performed on PCA-based geometries. The incompressible Navier–Stokes equations were solved with the following no-slip boundary conditions:

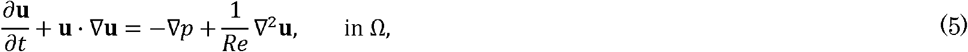

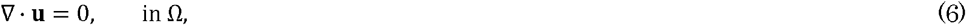

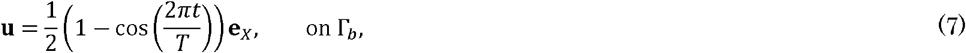

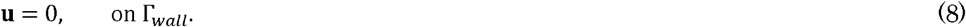

Here, **u** was the velocity vector, *p* was the pressure, and *Re* was the Reynolds number. In the present study, we focused on the feasibility of the proposed approach using data-driven synthetic geometry. The cavity flow–type velocity condition (7) was simply given in the vessel direction *X* on the aneurysm inlet surface Γ_*b*_, and the fixed wall condition (8) was applied on the aneurysm wall Γ_*wall*_. A pulsatile velocity was approximated by a cosine-bell wave form with a non-dimensional periodic time *T*. We set the parameters as *Re* = 100 and *T* =9.6.

The Cartesian grid–based CFD solver (Ii et al., 2017), which uses the pressure-projection method and boundary data immersion method (Weymouth and Yue, 2002), was applied. A uniform grid resolution of Δ*X =* Δ*Y =*Δ*Z=*3.0/52 ≈ 0.0577 was used, and the time resolution was set to Δ*t =*0.0004, resulting in a maximum Courant–Friedrichs–Lewy number of approximately 0.0069. The simulation was performed for five pulsatile cycles, and the results from the final cycle were used for analysis.

### 2.5 Hemodynamic metrics

Two hemodynamic metrics were evaluated in the current study: time-averaged WSS (TAWSS) and the oscillatory shear index (OSI). These metrics are widely used as fundamental metrics in hemodynamic analyses (Loly et al., 2025) and were defined as follows:

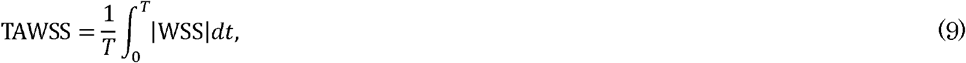

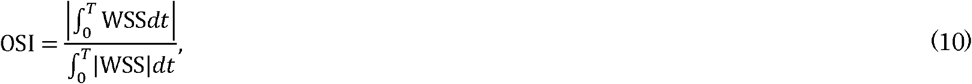

The WSS was calculated using the Cartesian-based method (Ii et al., 2017).

## 3. Results

### 3.1 PCA for patient-specific aneurysm geometries

Table 1 shows the singular values and contribution rates for respective principal components in PCA. The contribution rate of the first principal component was 51.91%, which was significantly higher than that of the second principal component (18.76%). The cumulative contribution rate exceeded 90% after the fourth principal component.

**Table 1.** Contribution of principal components in principal component analysis.

| Principal component | Singular value [-] | Contribution rate [%] |
| --- | --- | --- |
| 1 | 8.47 | 51.91 |
| 2 | 5.09 | 18.76 |
| 3 | 4.12 | 12.27 |
| 4 | 3.12 | 7.07 |
| 5 | 2.82 | 5.77 |
| 6 | 2.41 | 4.22 |
| 7 | 5.60E-15 | 2.27E-33 |

To examine how the truncation of principal components influences the reconstructed geometry, we varied the number of applied principal components *M*_*PC*_ ≤ *N* _*σ*_ in equation (3). As *N*_*PC*_ increased, the reconstructed geometry converged toward the original geometry (Fig. 3). At *M*_*PC*_ =4, where the cumulative contribution rate reached 90%, most samples reproduced the overall geometry well, although several exhibited reduced fidelities. S1 and S5 were accurately reconstructed with fewer than five components (*M*_*PC*_ < 5), whereas S4 required nearly the full set (*M*_*PC*_ =6) to recover its characteristic shape. For S3, although the overall shape was captured at relatively low *M*_*PC*_, a concave surface appeared around a distal region, and this feature was reproduced until at least *M*_*PC*_ = 6. By contrast, the characteristic shape of S6 was captured at *M*_*PC*_ = 1.

**Fig. 3.**
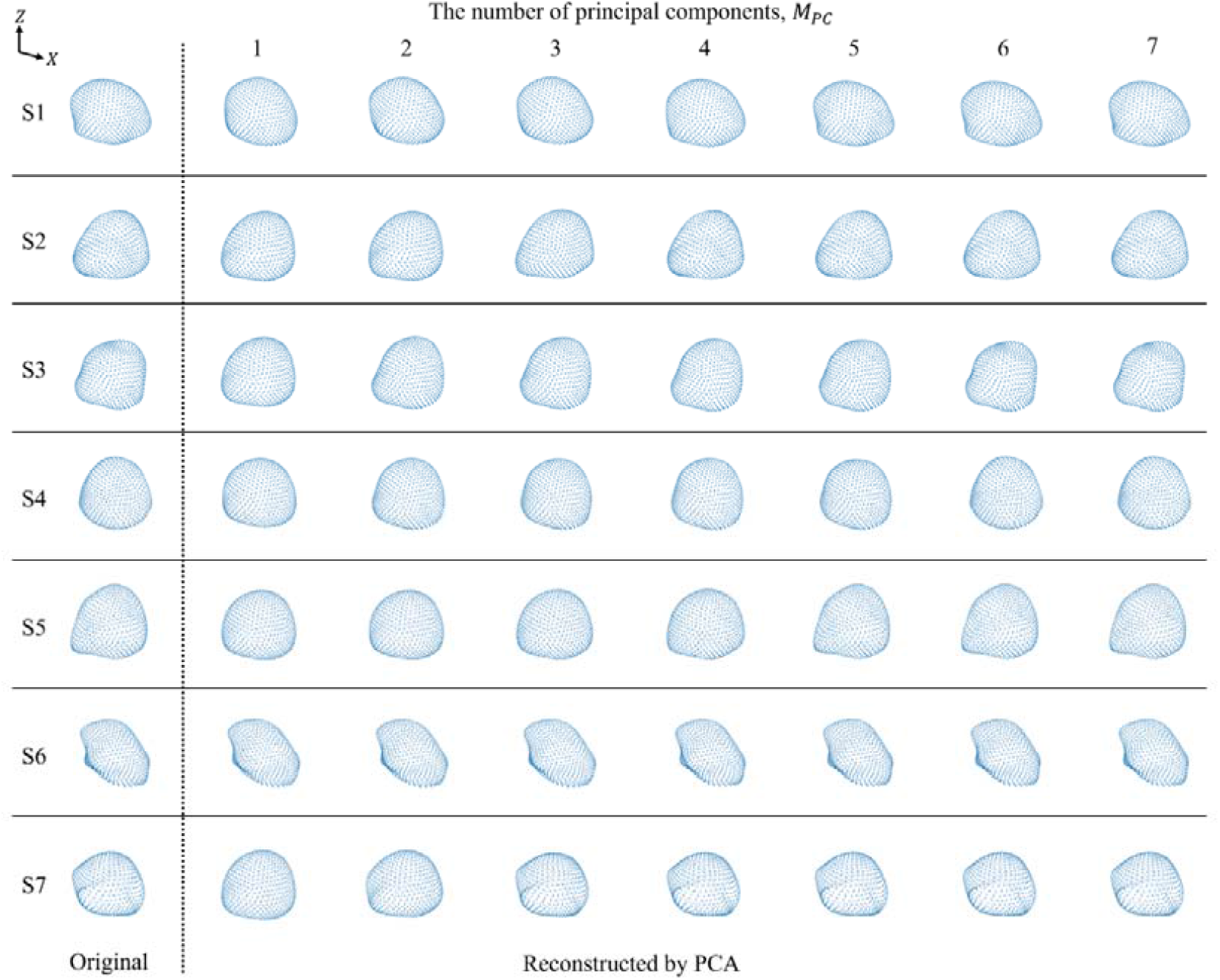
Contribution of the number of principal components *M*_*PC*_ in principal component analysis (PCA) for each sample (S1–S7). All the figures are shown with the same view position and angle.

Figure 4 summarizes the scores of principal components for all samples (S1–S7). S6 exhibited the smallest PCS1, and its PCS1 was relatively different from those of the other samples, which is consistent with the finding that the overall feature of S6 was unique in the present samples (Fig. 3). According to their similarity for principal components, similar geometrical features were observed in sets of {S2, S3}, {S4, S5}, and {S5, S7} for PCS1 and {S1, S7}, {S2, S5}, and {S3, S4} for PCS2. This geometrical similarity was also confirmed by the expansion up to PCS2 in Fig. 3.

**Fig. 4.**
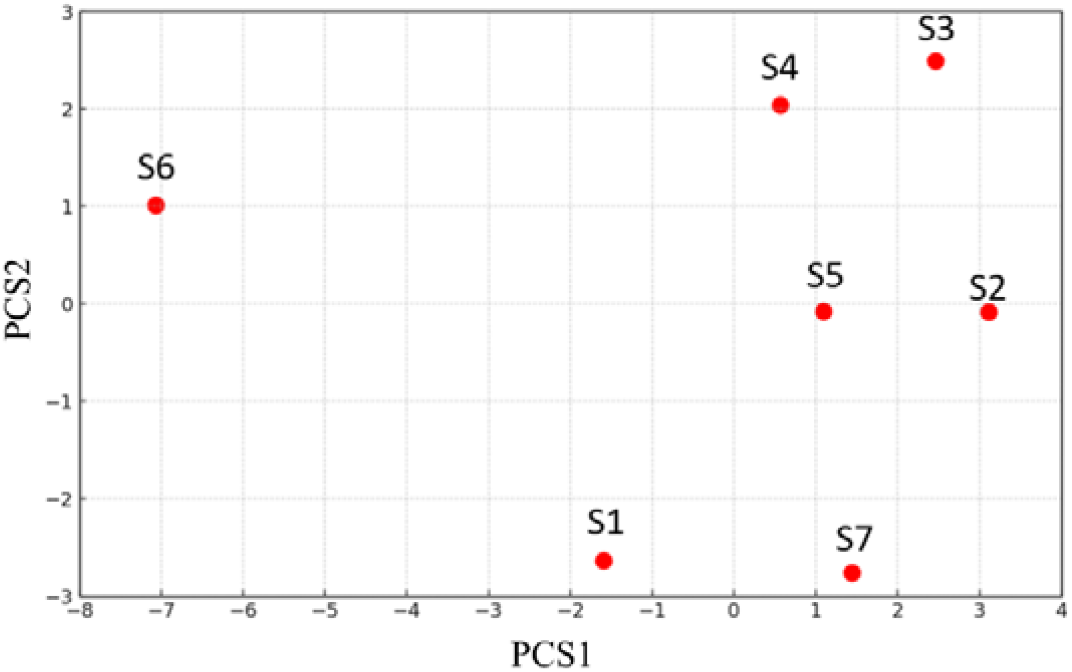
Plots of the first and second principal component scores (PCS1 and PCS2, respectively) for the samples (S1–S7).

### 3.2 PCA-based synthetic aneurysm geometries

We then investigated how major principal components can change aneurysm geometry. Here, we focused on aneurysm S5, whose PCS1 and PCS2 were within the range of those of the other samples. Figure 5 shows the shape changes of S5 for PCS1 and PCS2, respectively, where PCS were fixed to their own values. As PCS1 increased, the oblique shape in the *X* − *Z* plane transformed into a symmetric dome shape, and the inlet (neck) region became enlarged. This may be regarded as a change in the aspect ratio between the aneurysm height direction *Z* and the blood flow direction *X*. For PCS2, the aspect ratio changed between the blood flow direction *X* and its perpendicular direction *Y* on the aneurysm neck.

**Fig. 5.**
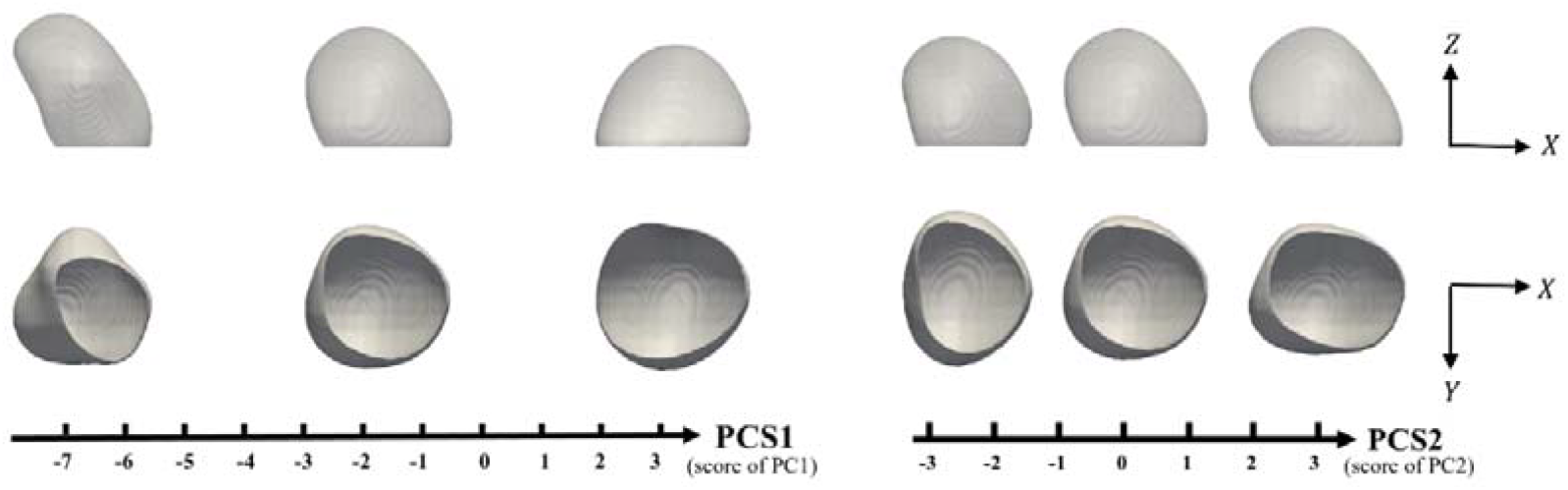
Effects of the first and second principal component scores (PCS1 and PCS2, respectively) on aneurysm geometry for S5.

We next generated nine synthetic geometries (A–I) by changing *α*_1_ and *α*_2_ (i.e., PCS1 and PCS2) for S5 using equation (4), where higher *α*_*k*_ for *k* ≥ 3 was fixed to those of S5. The scores were chosen using a 3 × 3 combination with *α*_1_ = {−7,− 2,3} and *α*_2_ = {−2.5, 0,2} (Fig. 6). In this regime, a similar tendency for geometric changes depending on PCS1 and PCS2 was observed.

**Fig. 6.**
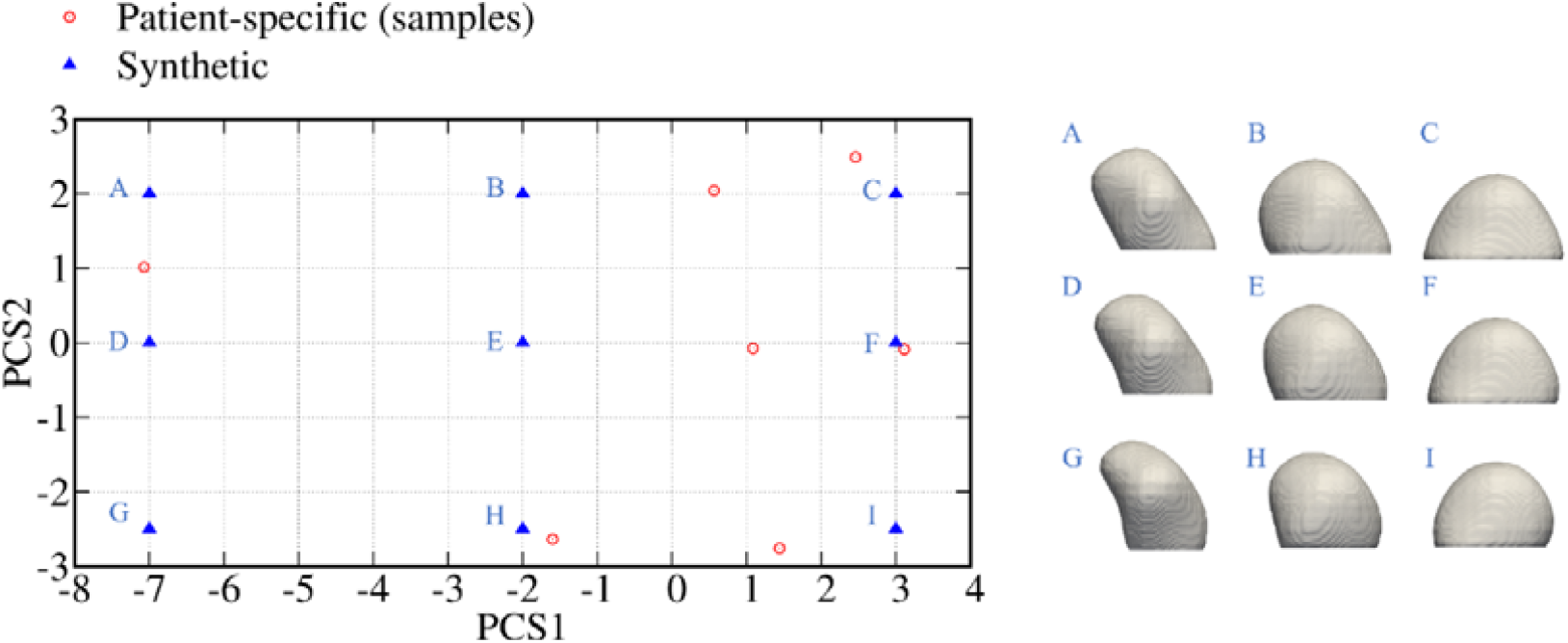
Principal component analysis–based synthetic aneurysm geometries for sample 5, which were generated by varying the first and second principal component scores (PCS1 and PCS2, respectively).

### 3.3 CFD for PCA-based synthetic aneurysm geometries

The CFD results for PCA-based aneurysm models A–I are summarized in Fig.□7, in which the velocity fields at the peak inlet velocity phase are presented. Across all geometries, a consistent circulation pattern was observed, which was characterized by a vortex structure that formed near the aneurysm inlet. The velocity markedly decreased near the dome apex, and this reduction was particularly pronounced in geometries A, D, and G, which exhibited relatively greater dome heights associated with low PCS1 values.

**Fig. 7.**
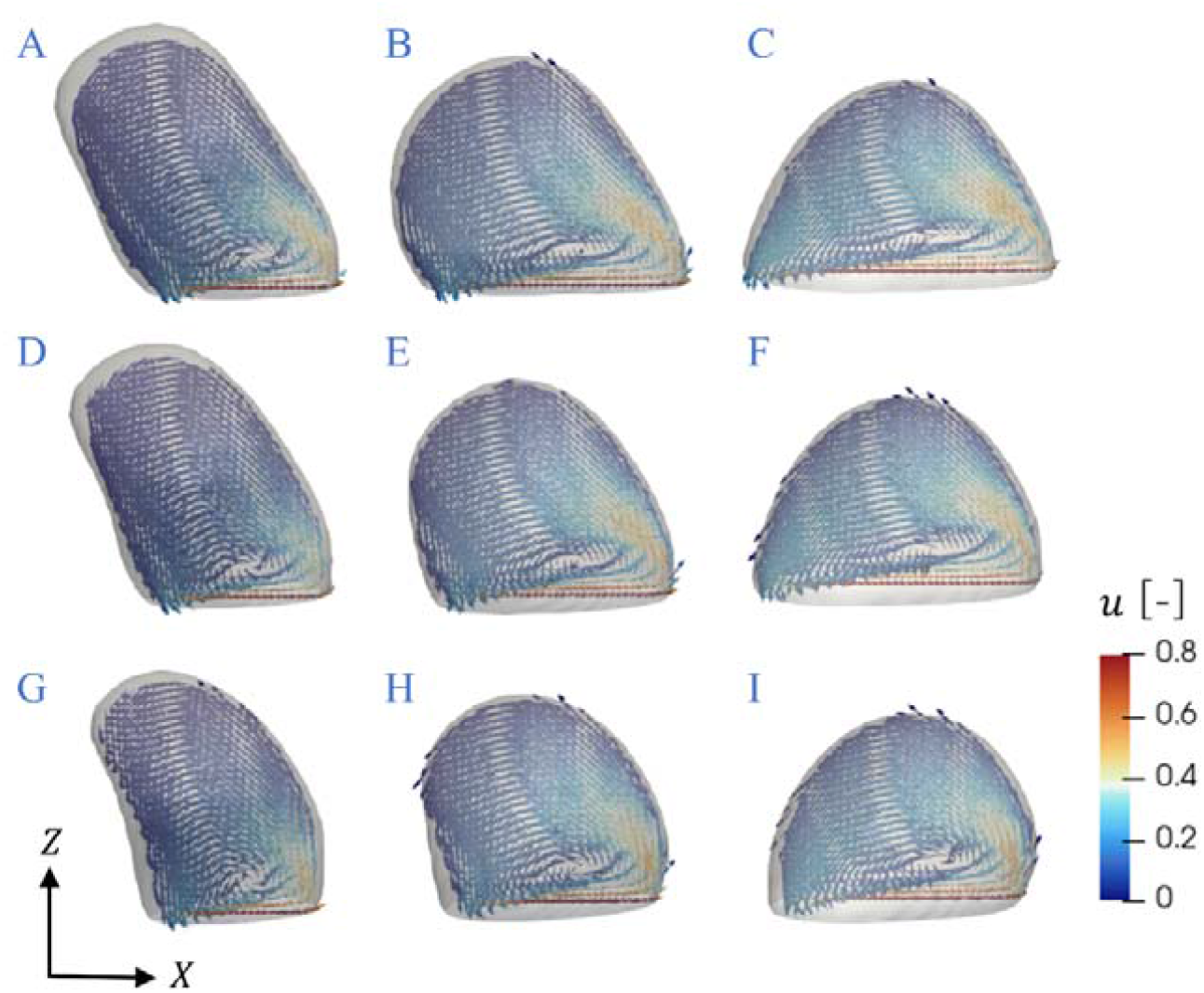
Velocity fields obtained from computational fluid dynamic simulations using principal component analysis–based aneurysm models. (A–I) Flow patterns at the peak inlet velocity phase.

Figure 8(A), (B) presents the spatially averaged values of TAWSS and OSI (mean TAWSS and OSI) as a function of PCS1. As PCS1 increased, the mean TAWSS increased, whereas the mean OSI decreased. This tendency became more pronounced when the PCS2 was larger, particularly in the case of the mean OSI, for which the effect was substantial. Figure 8(C) shows the spatial distribution of WSS for geometries A, B, and C at the peak inlet velocity phase. As PCS1 increased, the aneurysmal dome height decreased; this resulted in not only an overall elevation of WSS across the wall but also a marked increase at regions in which the upward-circulating flow from the aneurysm bottom impinged, as well as along the lateral surface near the vorticity center.

**Fig. 8.**
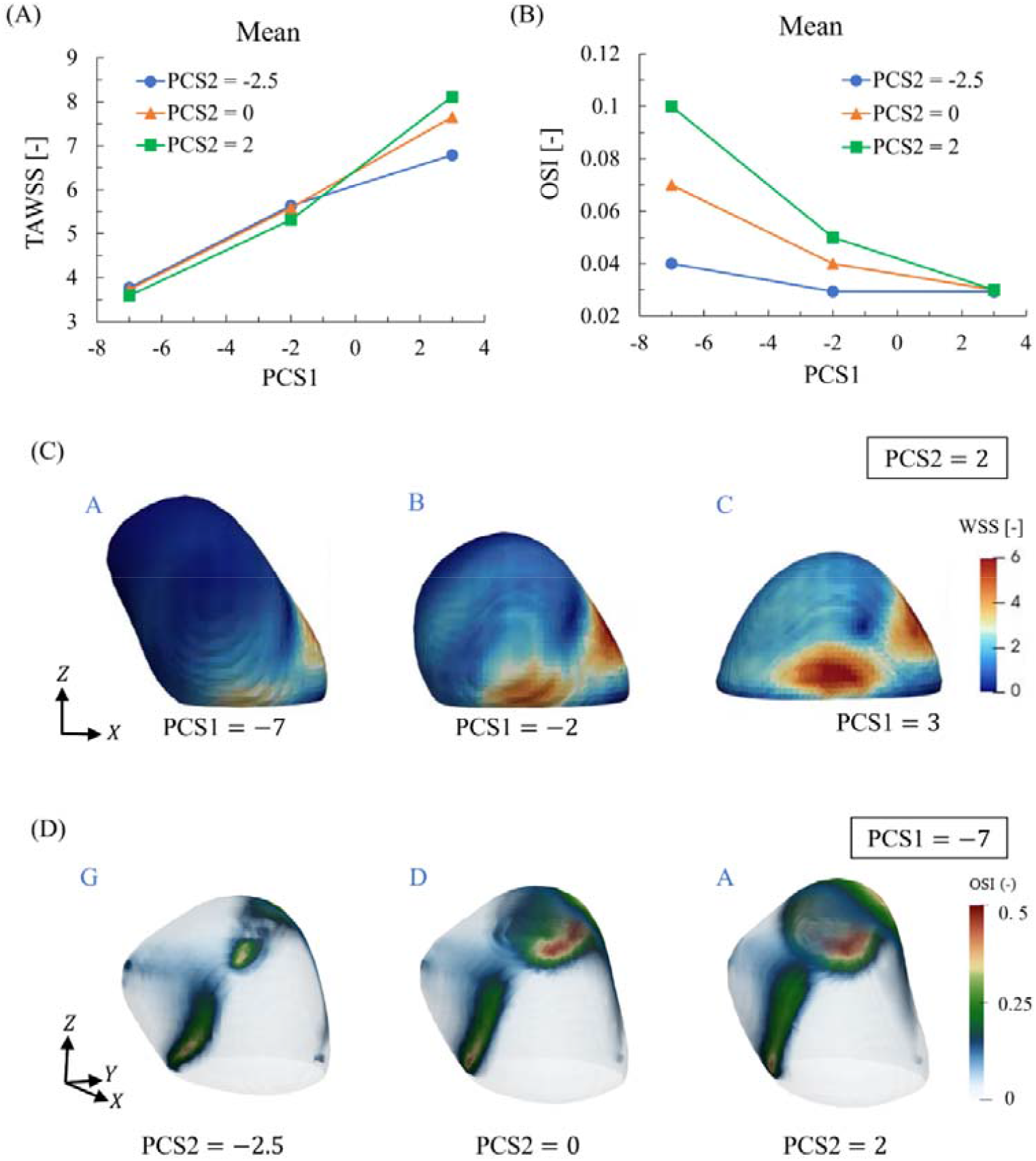
The mean time-averaged wall shear stress (TAWSS) (A) and oscillatory shear index (OSI) (B) as a function of the first principal component score (PCS1) for different second principal component score (PCS2) values. Panel (C) shows the spatial distributions of wall shear stress for geometries A, B, and C at the peak inlet velocity phase. Panel (D) shows the distribution of OSI for geometries G, D, and A.

The spatial distributions of OSI for each PCS2 under the condition of PCS1 = −7 are presented in Fig. 8(D). In addition to the line-shaped region of elevated OSI from the aneurysm inlet on the surface perpendicular to the *Y*-axis, the aneurysm dome exhibited two bump-like protrusions at PCS1 = −7, and one of these regions showed a pronounced increase in OSI. This tendency became more pronounced with a large PCS2.

## 4. Discussion

For the seven patient samples, PCA revealed that the cumulative contribution rate exceeded 90% by the fourth principal component. In most cases, the overall geometry was reproduced well at this level; however, some samples required additional principal components to capture their local geometric features. This finding suggests substantial variability across individuals. In general, a cumulative contribution rate of 90% indicates that most of the information in the original dataset is retained, thus serving as a metric for dimensionality reduction. However, fine geometric features represented by higher-order principal components, such as local protrusions and inlet morphology, can have a substantial effect on hemodynamics. Previous studies of vascular shape analysis using PCA have also demonstrated that higher-order principal components are required depending on the morphological features of interest (Wiputra et al., 2023; Eulzer and Lawonn, 2025). Together, these results suggest that higher-order principal components must be considered for generating synthetic datasets when local hemodynamics are being evaluated.

Eulzer and Lawonn (2025) mapped three-dimensional aneurysm geometries onto a two-dimensional parametric domain and performed PCA on surface points (of mesh) attached to that domain. However, parametric representations can introduce singularities near the poles and the non-uniform spatial resolution of surface elements, which may affect statistical shape analysis and the interpretability of principal modes. By contrast, our approach matched the nodes of a template mesh to each aneurysm surface via CPD-based non-rigid registration in the physical domain. This ensures uniform and well-distributed surface sampling before PCA, thereby mitigating biases arising from resolution non-uniformity.

In the present study, S6 exhibited a noticeably different overall morphology compared with the other samples, and it was also located far from the other samples in PCS with respect to both PCS1 and PCS2. Although the present evaluation focused only on PCS1 and PCS2, morphological differences among the samples appeared to be related to the proximity of their scores in this reduced space. Our results suggest that aneurysm geometries may be classified by using the proximity of PCS as norms of their differences. Although the limited number of samples in the current study prevented a sufficiently robust classification, increasing the dataset size in future work may enable more reliable clustering, which might facilitate the association of aneurysmal hemodynamics with a smaller set of explanatory variables.

By varying PCS1 and PCS2 arbitrarily and examining their effects on aneurysm geometry, we revealed that PCS1 primarily altered the height and width of an aneurysm, whereas PCS2 changed its width and depth. In the present study, the volume of each sample was normalized, and the aspect ratios of the aneurysms across samples were therefore likely evaluated as the dominant principal component. Particularly, there was relatively large variability in aneurysm height among the present samples. Interestingly, a previous PCA-based study on geometrical analysis using different aneurysm datasets (Eulzer and Lawonn, 2025) reported that the dominant principal components (up to the second one) produce deformation patterns that are similar to those observed in the present study. Although the number of samples in our study was extremely limited compared with that in the prior work, the similar deformation tendencies that were obtained reinforces the notion that the primary source of morphological variability in aneurysms is the aspect ratio. The major shape variations that were identified in those samples—namely, changes in the aspect ratio—can be identified quantitatively by applying PCA. Although such variations might be qualitatively inferred through visual inspection, it would be difficult to translate these insights into a practical framework for generating arbitrary geometries from real geometries. By contrast, the PCA approach may allow the generation of arbitrary geometries in a manner that preserves geometric naturalness relative to the original aneurysm geometries.

In the CFD analysis, a boundary velocity was applied in the blood flow direction, meaning that the overall circulation pattern was similar to a cavity flow. This circulation pattern depended little on PCS2 and was primarily influenced by PCS1, which affected aneurysm height. The evaluation of WSS revealed that aneurysm shapes with lower heights and a more symmetric dome geometry (i.e., larger PCS1) exhibited higher TAWSS and a dependence on PCS2. By contrast, shapes with greater aneurysm heights and an upstream-oblique configuration (i.e., smaller PCS1) showed higher OSI and a strong dependence on PCS2. For the latter case, the increase in PCS2 enlarged the aspect ratio in the *Y*-direction, which was perpendicular to the flow direction *X*, and consequently displaced the dome region containing one of the bumps farther from the aneurysm inlet. This may increase the flow fluctuation near the wall relative to the average flow around the bump, which might in turn lead to an increase in OSI. In aneurysmal remodeling, previous studies have demonstrated that OSI increases in bleb regions (Nordahl et al., 2021; Cornelissen et al., 2022), which is consistent with the findings of the present study. In our analysis, the ranges of PCS1 and PCS2 were constrained to the minimum and maximum values that were observed in the real samples to ensure that the generated geometries remained within a physiologically relevant domain. However, by extrapolating these ranges or incorporating post-growth aneurysm geometries into the dataset, it may be possible to further elucidate the relationship between aneurysm growth and hemodynamic factors.

Recent advances in deep learning for hemodynamic assessment as an alternative to computational mechanics have been remarkable (Morales et al., 2020; Yuhn et al., 2022; Taebi, 2022; Du et al., 2022; Djuansjah et al., 2025; Liao et al., 2025), and such approaches have also been applied to cerebral aneurysms (Li et al., 2021; Rajhi et al., 2025). In these studies, CFD analyses under diverse conditions were performed using synthetic geometries, and the resulting datasets are essential as training data (Faisal et al., 2025; Li et al., 2025). Although no prior studies have specifically targeted cerebral aneurysms, the proposed method is expected to provide a useful technique for generating such training datasets.

There were several limitations of the present study. First, the CFD analysis used a cavity flow condition in which no velocity components were applied in the *Y* – or *Z*-directions as the boundary condition at the aneurysm neck. In actual blood flow environments, however, the inflow into an aneurysm is accompanied by diverse velocity profiles and inflow angles (Cebral et al., 2005; Xiang et al., 2010). Future analyses should therefore consider a broader range of inlet velocity conditions. Second, synthetic geometries were generated only by varying the PCSs up to the second principal component, and the relationships between higher-order PCSs and hemodynamic factors were not investigated. This limitation mainly stems from the need to suppress the increase in computational cost that is associated with the combinatorial growth of simulations. In future work, it would be desirable to adopt more efficient approaches for covering the geometry space, such as adjusting the number of samples for each PCS based on their contribution ratios. Third, the dataset consisted solely of aneurysms in the right internal carotid artery and did not include aneurysms in other vascular regions, nor did it account for more complex geometries such as those with blebs (Nordahl et al., 2021; Cornelissen et al., 2022). Incorporating such anatomical variations into CFD analyses would not only facilitate a more comprehensive understanding of the relationship between aneurysm geometry and hemodynamic factors but also contribute to the development of advanced surrogate models with reduced computational costs. Finally, the hemodynamic indices were limited to TAWSS and OSI; the use of more indices would help to provide a comprehensive understanding of the relationship of hemodynamics with physiological aspects of aneurysm growth.

## 5. Conclusion

Here, we proposed a systematic approach for CFD using data-driven synthetic geometries to evaluate the hemodynamics of cerebral aneurysms. The PCA was applied to point clouds of cerebral aneurysms that were derived from patient-specific data, and novel aneurysm geometries were generated by varying PCAs within an arbitrary range. By performing CFD using the synthetic geometries, we were able to associate variations in major principal components with corresponding changes in intra-aneurysmal flow dynamics, WSS, and associated hemodynamic metrics (mean TAWSS and OSI). Together, this enabled us to quantitatively assess the relationship between aneurysm geometry and hemodynamic factors.

## Data Availability

All data produced in the present study are available upon reasonable request to the authors

## Acknowledgments

The authors thank Marie Oshima (The University of Tokyo), Shigeo Wada (Osaka University), Tomohiro Otani (Osaka University), and Hiroki Suzuki (Institute of Science Tokyo) for fruitful discussions on patient-specific analysis and principal component analysis. This work was supported by the MEXT Program for Promoting Researches on the Supercomputer Fugaku (Development of human digital twins for cerebral circulation using Fugaku, JPMXP1020230118) and used computational resources of the supercomputer Fugaku, provided by the RIKEN Center for Computational Science (project ID: hp230208, hp240220, and hp250236). Some computations were also carried out using the supercomputer “Flow” at the Information Technology Center, Nagoya University. This work was also supported by JSPS KAKENHI (Grant Numbers JP22H00190 and JP24K02557). We thank Bronwen Gardner, PhD, from Edanz (https://jp.edanz.com/ac) for editing a draft of this manuscript.

